# Deep Learning for Multi-Label Disease Classification of Retinal Images: Insights from Brazilian Data for AI Development in Lower-Middle Income Countries

**DOI:** 10.1101/2024.02.12.24302676

**Authors:** Dewi S.W. Gould, Jenny Yang, David A. Clifton

## Abstract

Retinal fundus imaging is a powerful tool for disease screening and diagnosis in opthalmology. With the advent of machine learning and artificial intelligence, in particular modern computer vision classification algorithms, there is broad scope for technology to improve accuracy, increase accessibility and reduce cost in these processes. In this paper we present the first deep learning model trained on the first Brazilian multi-label opthalmological datatset. We train a multi-label classifier using over 16,000 clinically-labelled fundus images. Across a range of 13 retinal diseases, we obtain frequency-weighted AUC and F1 scores of **0.92** and **0.70** respectively. Our work establishes a baseline model on this new dataset and furthermore demonstrates the applicability and power of artificial intelligence approaches to retinal fundus disease diagnosis in under-represented populations.

## 1 Introduction

The field of medical imaging is growing at an exponential rate, resulting in a vast amount data that necessitates the expertise of medical professionals for interpretation and triaging. Across various clinical specialties, there exists a shortage of such expertise, leading to delays in diagnosis and referrals. Machine learning (ML) and artificial intelligence (AI) algorithms offer the potential to deliver healthcare solutions that are innovative, efficient, cost-effective, and readily accessible: ultimately improving the diagnosis and treatment of medical conditions on a global level. In the field of ophthalmology, AI holds substantial promise, particularly in the domain of computer vision (CV) classification algorithms [1]. This promise is exemplified by the existence of FDA-approved devices designed for diabetic retinopathy screening [2, 3].

In ophthalmology additional imaging procedures like retinal fundus images, optical coherence tomography, corneal topography, visual field tests, and anterior segment photos of the eye are crucial for screening, diagnosing, and continuously monitoring diseases. However, the widespread availability of these images has not been matched by an equal availability of clinical experts capable of interpreting the scans and referring patients to appropriate care pathways [2, 4]. This problem is further compounded by the increasing incidence of sight-threatening diseases, where retinal imaging is the primary means of assessment.

Although middle to high-income countries (MHICs) often have the resources to manage extensive image datasets and embrace cutting-edge AI technologies, implementing healthcare AI in hospitals within low- and middle-income countries (LMICs) can pose significant obstacles. In the field of ophthalmology, LMICs are confronted with a growing disparity between the quantity of ophthalmologists and the size of their populations [5]. This inequality is accentuated by the fact that two-thirds of the global ophthalmologist workforce is concentrated in only seventeen countries [4].

The availability of data is a crucial factor in AI model development. Currently, the majority of retinal datasets originate from MHICs. Thus, these datasets lack comprehensive representation of demographic diversity and comorbidities, with many predominantly focusing on diabetic retinopathy patients. This existing data imbalance can lead to biased and potentially harmful outcomes when these models are applied to populations in LMICs [5, 6].

Retinal fundus imaging provides a relatively cheap and acccessible tool for disease diagnosis [7]. The exploration of integrating artificial intelligence tools into this procedure has shown encouraging outcomes [8], with many existing works focusing on binary or small-number multi-label disease classification [9–12]. Furthermore, there have been various works extending this to more comprehensive multi-label disease classification [13–17].

In this study, we aim to develop a multi-label classifier to automatically label a range of retinal fundus diseases, using a dataset collected from a region so-far not represented in the literature (Latin America, and in particular Brazil).

In section 2 we discuss our methods, including details of the dataset used, our model and details of its training. In section 3 we present our results, and in sections 4 and 5 we have a discussion and conclusions.

## 2 Methods

### 2.1 Dataset

In this study, we use the Brazilian Multilabel Ophthalmological Dataset (BRSET) [18], which has been made available on PhysioNet [19]. See section 5 for information on accessibility. This dataset consists of 16,266 images from 8,524 Brazilian patients. This project was approved by the São Paulo Federal University - UNIFESP institutional review board (CAAE 33842220.7.0000.5505). The 8,524 patients represented in this dataset were collected from three ophthalmological centers. For each patient, only one paired exam focused on the macula is included.

The retinal photographs were taken using a Nikon NF505 camera (Tokyo, Japan), and a Canon CR-2 camera (Canon Inc., Melville, NY, USA). These were captured by non-medical professionals who had received prior training in pharmacological mydriasis.

The final dataset includes only fovea-centered images with both temporal retinal vascular arcades visible, as well as at least one disc diameter of retina visible on the nasal side of the optic disc. Furthermore, non-retinal images and fluorescein angiogram photos were excluded. These images were captured at a 45-degree angle and were centered on the optic disc. See Figure 1 for an example image.

**Fig. 1.**
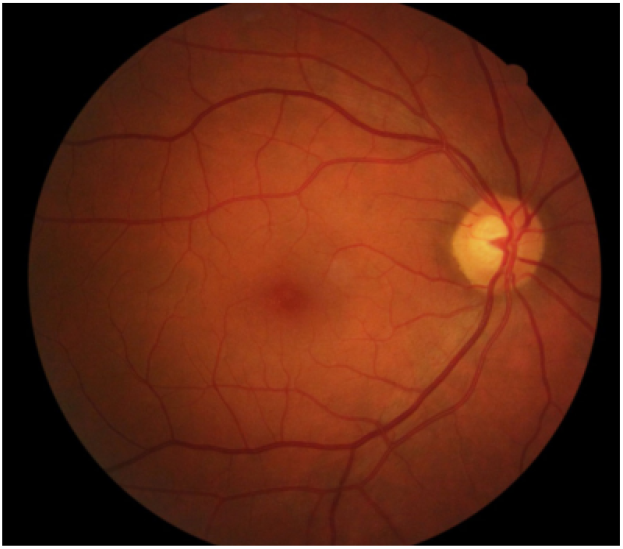
An example of a retinal fundus image in the dataset.

#### Dataset Preparation

In all color fundus images, all file identification, sensitive information (such as patient identity and examination date), and headers were removed. No preprocessing procedures were applied to the images after they were exported directly to JPEG format. It’s important to note that all images were captured with a focus on the macula, ensuring a consistent viewpoint.

#### Supplementary Features

Supplementary information related to retinal labeling includes details about the type of retinal camera device used, patients’ nationality, age (in years), gender, medical history, insulin usage, and the duration of diabetes. These demographic and medical characteristics were extracted from electronic medical records, which were based on self-reported medical history. 16% of the patients have diabetes, and the average age of the patient group is 57 years old.

With a focus on establishing the widest possible applicability of our model, we do not incorporate these features in the training of our deep learning model: we instead *only* use the images themselves.

#### Image Quality

The dataset contains various features describing image quality. A label of “satisfactory/ unsatisfactory” is assigned in each of the following areas: image focus, illumination, image field and artefacts. An image is assigned a quality score of “Inadequate” if any of these are unsatisfactory. In the entire dataset there are 1987 images (12%) of this type.

Since we are interested in building a model with optimal generalizability to realworld data, we include these images in our training and testing datasets. However, in section 3.2.4 we run an experiment where we remove all “Inadequate” images from the dataset for comparison.

#### 2.1.1 Clinical Labeling

Every image underwent labeling by an ophthalmologist with specialization in retinal conditions, and the BREST research team formulated the criteria for the labeling process. Under the anatomical classification, the retinal optic disc, retinal vessel, and macula features were categorized as either “Normal” or “abnormal”.

Each image was evaluated for a list of pathological conditions which we summarize, along with their respective number of occurences, in Table 1. Note that “Other” describes a collection of retinal diseases not included in the initial list, whilst “Normal” is for any images not falling into these categories. For completeness, we also summarize the number of occurences of eyes labelled with more than one condition in Table 2. For the classification of diabetic retinopathy, both the International Clinic Diabetic Retinopathy (ICDR) grading system and the Scottish Diabetic Retinopathy Grading (SDRG) system were utilized.

**Table 1.**
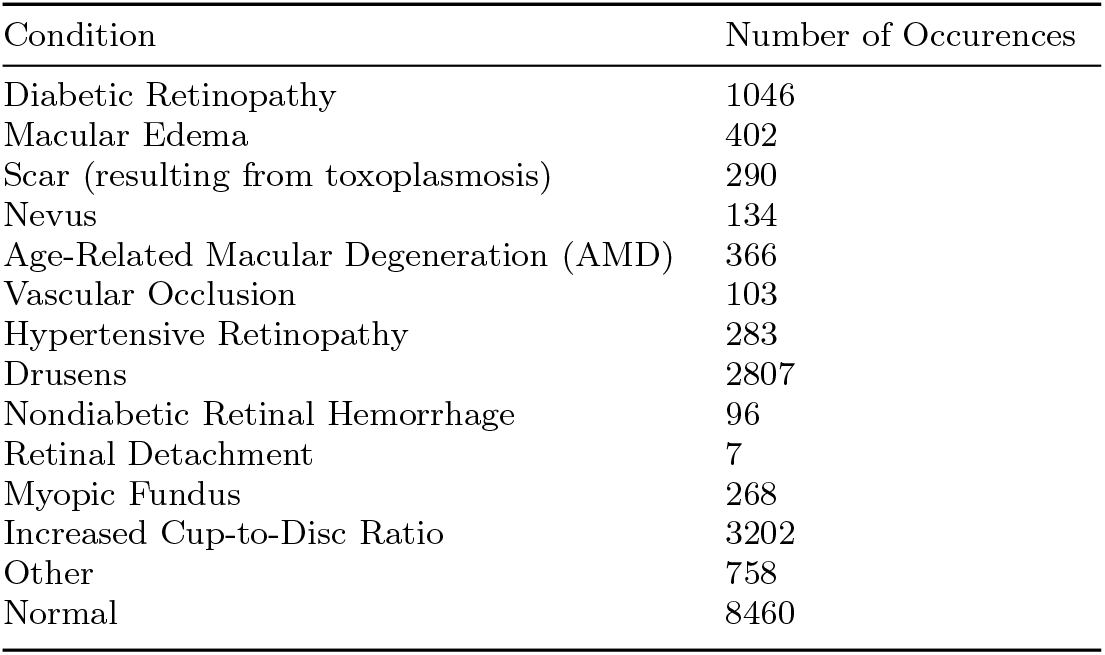
Number of occurences of pathological conditions in dataset.

**Table 2.** Number of images with multiple conditions.

| Number of Unique Conditions | Number of Occurrences |
| --- | --- |
| 0 | 8460 |
| 1 | 6017 |
| 2 | 1622 |
| 3 | 167 |

**Table 3.** Dataset split by camera type.

| Camera | Number of Images |
| --- | --- |
| Canon | 10592 |
| Nikon | 5674 |

The dataset encompasses the entire spectrum of medical retina patients who sought care and underwent treatment at the three ophthalmological centers. While the specific pathological labels were selected by the specialist othalmologist and the BREST research team, the framework introduced in this study can be adapted to suit the needs of other healthcare systems, including those with distinct classification requirements.

### 2.2 Model Development

#### 2.2.1 Preprocessing

The dataset contains 16,266 images in JPEG format. In the pre-processing stage, these image inputs were standardized to be of consistent size. During training, various augmentation procedures were employed to increase dataset diversity. These include a random choice of the following: rotation by angle *θ* ∈ [−30^*?*^, 30^*?*^], re-scaling by a parameter *C* ∈ [1.0, 1.25], image warping, and horizontal flipping. These steps were not applied to the validation or test sets. Furthermore, all pixels values were normalized to have zero mean and unit variance.

##### Addressing Class Imbalance

The distribution of the 13 classes in the dataset is highly imbalanced (see Table 1). There are various approaches to dealing with imbalanced data including under/over-sampling and ensemble methods. In this work we experiment with various strategies to account for this effect, see Section 3.2.2.

#### 2.2.2 Training Architecture

##### Deep Learning in Image Classification

Deep learning for image classification involves training neural networks, most often convolutional neural networks (CNNs), to extract key features from input pixels relevant for a classification problem. CNNs are special neural networks containing so-called *convolutional* layers. These layers allow the network to learn spatial features and correlations of an input image.

##### ResNet-50

Residual Network 50 (ResNet-50)[20] is a powerful deep learning model using for computer vision tasks. Concretely, it is a convolutional neural network with 50 layers. In this project we begin our experiments with the pre-trained ResNet-50 model, the weights of which are obtained after training on the ImageNet [21] database.

The fundamental element of the ResNet-50 model is the residual learning block. This is a block of several layers which, for a given input *x*, produces a standard non-linear output ℱ(*x*) *plus* the input *x*:

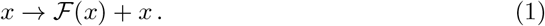

This is implemented by the addition of so-called “shortcut connections”, decorated with the identity map, which connect the input and output without passing through the non-linear layers. Adding these components to a CNN provides a way to address existing issues with training error increasing with network depth.

##### Full Architecture

Our full model architecture includes the ResNet-50 model and a dense final layer constructed with sigmoid activiation function and *N*_classes_ (= number of classes) outputs^1^.

##### Parameters

For our purposes, we remove the pre-trained fully-connected layer of the baseline ResNet model and replace it with our own. We use an Adam optimizer and binary cross entropy loss function when training our models. For all experiments we use a 60 − 20 − 20 split between training, validation and testing sets respectively.

##### Implementation Details

Throughout this project we used TensorFlow 2.12.1 [22] with Python 3.9.

##### Optimizing Thresholds

A crucial point to note is that the output of our model is *N*_classes_ *continuous* predictions (probabilities ranging from 0 to 1). In order to turn these continuous variables into disease predictions we must place *thresholds* on these outputs. In other words, we have the freedom to select *N*_classes_ thresholds which will convert the continuous outputs to binary predictions for each disease. The default choice for these thresholds is always 0.5.

For a class *i*, we denote the threshold for the corresponding sigmoid activiation function *τ*_*i*_. One way of choosing a tentative set 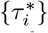 of “optimal” values for these, which we employ in various experiments in this work, is the following. The set 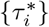 can be dynamically chosen via grid search to be those which maximise the individual F1 scores:

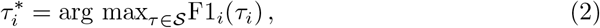

where *𝒮* is some set of values between 0 and 1. In this work, we chose *𝒮* = {0, 0.04, 0.08, …, 1}.

##### Sensitivity Analysis

As explained above, the dataset contains images taken on two different types of camera, Nikon NF505 and Canon CR-2. We will refer to these as “Nikon” and “Canon” respectively from now on for notational convenience. We performed two experiments, where in each we trained our model on images taken with only one type of camera. We then tested the power of these models to generalize by testing on the images taken with the other type. The results of this analysis are contained in Section 3.2.3.

## 3 Results

In this section we analyze various models in terms of their classification performance on the 13 retinal conditions.

### 3.1 Metrics

Using the abbreviations TP/FP/TN/FN for true positive, false positive, true negative and false negative rates respectively, the fundamental metrics we will use for performance within each class, are

- Accuracy: This is computed as the percentage of correct predictions.
- Sensitivity/ Recall: This is computed as 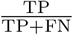.
- Specificity: This is computed as 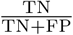.
- PPV/ Precision: This is computed as 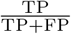.
- NPV: This is computed as 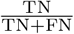.
- F1-score: This is the *harmonic* mean of Precision and Recall:

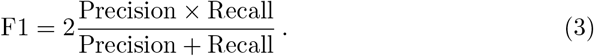
- Area under the ROC curve score (AUC): This is the area under the curve of true-positive rate against false-positive rate plotted at various thresholds. The AUC is a threshold-independent metric and is particularly useful in the case of imbalanced datasets like ours.

For class-wide performance, we will use the *frequency-weighted* f1-score, computed via

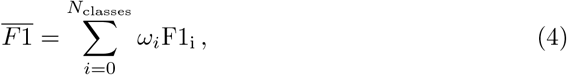

where *ω*_*i*_ is the fraction of the test set which belongs to class *i*. We will also denote by 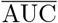 the frequency-weighted average AUC metric.

### 3.2 Performance

#### 3.2.1 Baseline Model

After experimenting with various numbers of epochs and batch sizes, the best performing model was with parameters: 6 epochs and batch size 8. We explain this experimentation procedure in more detail in Section 4. Having established these baseline parameters, we then incorporated the validation set into the training set to allow the model to learn from more data. The performance of this model is given in Table 4. In this Table, we have dynamically chosen optimal threshold parameters, via the method described around (2). See Table A2 for the same model with the default threshold values (0.5) and Table A1 for the model without the validation set incorporated.

**Table 4.**
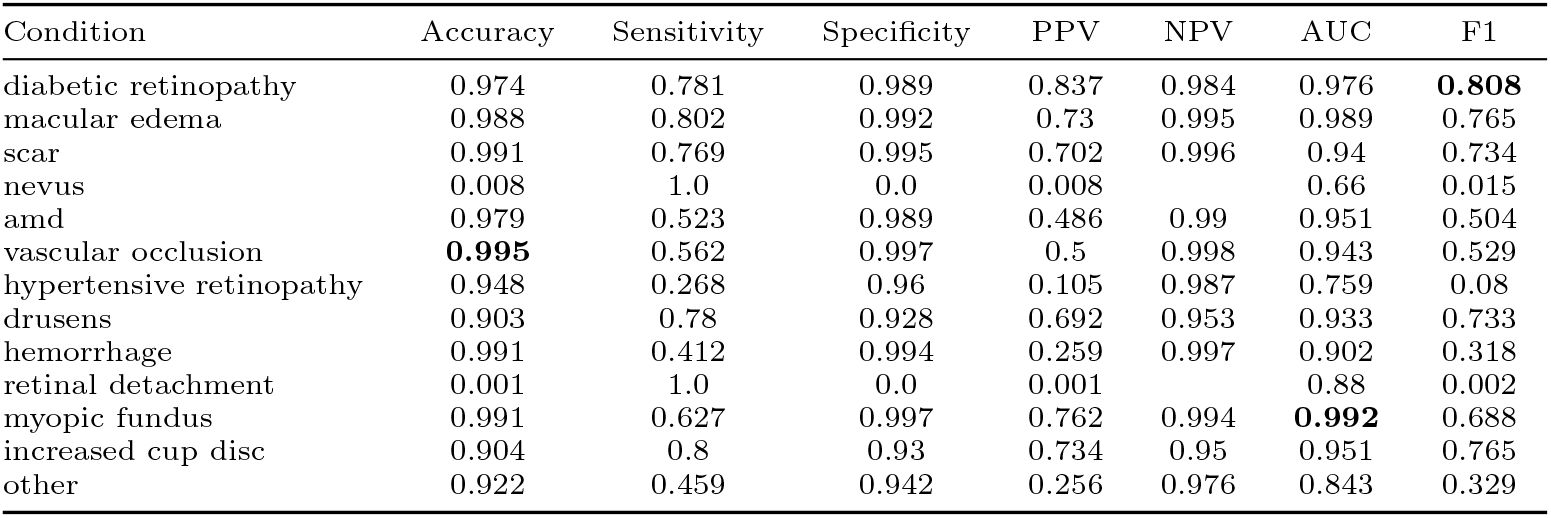
Performance metrics for various eye conditions. Trained for 6 epochs on the training and validation sets. This model has 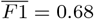 and 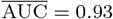. For clarity we have highlighted the row with the best accuracy, AUC and F1 score with boldface in each table.

| Condition | Accuracy | Sensitivity | Specificity | PPV | NPV | AUC | F1 |
| --- | --- | --- | --- | --- | --- | --- | --- |
| diabetic retinopathy | 0.974 | 0.781 | 0.989 | 0.837 | 0.984 | 0.976 | **0.808** |
| macular edema | 0.988 | 0.802 | 0.992 | 0.73 | 0.995 | 0.989 | 0.765 |
| scar | 0.991 | 0.769 | 0.995 | 0.702 | 0.996 | 0.94 | 0.734 |
| nevus | 0.008 | 1.0 | 0.0 | 0.008 |  | 0.66 | 0.015 |
| amd | 0.979 | 0.523 | 0.989 | 0.486 | 0.99 | 0.951 | 0.504 |
| vascular occlusion | **0.995** | 0.562 | 0.997 | 0.5 | 0.998 | 0.943 | 0.529 |
| hypertensive retinopathy | 0.948 | 0.268 | 0.96 | 0.105 | 0.987 | 0.759 | 0.08 |
| drusens | 0.903 | 0.78 | 0.928 | 0.692 | 0.953 | 0.933 | 0.733 |
| hemorrhage | 0.991 | 0.412 | 0.994 | 0.259 | 0.997 | 0.902 | 0.318 |
| retinal detachment | 0.001 | 1.0 | 0.0 | 0.001 |  | 0.88 | 0.002 |
| myopic fundus | 0.991 | 0.627 | 0.997 | 0.762 | 0.994 | **0.992** | 0.688 |
| increased cup disc | 0.904 | 0.8 | 0.93 | 0.734 | 0.95 | 0.951 | 0.765 |
| other | 0.922 | 0.459 | 0.942 | 0.256 | 0.976 | 0.843 | 0.329 |

#### 3.2.2 Class Imbalance

We performed under-sampling on the majority class (“Normal”) by removing 5000 of these images from the dataset to address the imbalance between this class and all others. The results of this training are in Table 5. The performance of this model is marginally better than the baseline in Table A2.

**Table 5.** Performance metrics for various eye conditions. 5000 random “Normal” images removed from dataset before training. Frequency-weighted F1 and AUC scores are 0????????70 and 0????????92 respectively.

| Condition | Accuracy | Sensitivity | Specificity | PPV | NPV | AUC | F1 |
| --- | --- | --- | --- | --- | --- | --- | --- |
| diabetic retinopathy | 0.971 | 0.784 | 0.988 | 0.861 | 0.98 | 0.972 | **0.821** |
| macular edema | 0.983 | 0.738 | 0.992 | 0.776 | 0.99 | 0.959 | 0.756 |
| scar | 0.985 | 0.769 | 0.992 | 0.735 | 0.993 | 0.933 | 0.752 |
| nevus | 0.987 | 0.043 | 0.996 | 0.111 | 0.99 | 0.671 | 0.062 |
| amd | 0.954 | 0.541 | 0.968 | 0.364 | 0.984 | 0.937 | 0.435 |
| vascular occlusion | 0.984 | 0.652 | 0.987 | 0.349 | 0.996 | 0.971 | 0.455 |
| hypertensive retinopathy | 0.973 | 0.278 | 0.99 | 0.405 | 0.982 | 0.84 | 0.33 |
| drusens | 0.871 | 0.803 | 0.895 | 0.727 | 0.928 | 0.92 | 0.763 |
| hemorrhage | **0.991** | 0.211 | 0.997 | 0.4 | 0.993 | 0.88 | 0.276 |
| retinal detachment | 0.001 | 1.0 | 0.0 | 0.001 |  | 0.276 | 0.002 |
| myopic fundus | 0.986 | 0.709 | 0.993 | 0.722 | 0.993 | **0.984** | 0.716 |
| increased cup disc | 0.874 | 0.811 | 0.9 | 0.77 | 0.92 | 0.94 | 0.79 |
| other | 0.889 | 0.45 | 0.919 | 0.268 | 0.962 | 0.795 | 0.336 |

We also tried amalgamating some of the poorer performing classes within the “Other” column. For the experiment in Table 6 we combined the diseases retinal detachment, hemorrhage, nevus and hypertensive retinopathy in this way.

**Table 6.** Performance metrics for various eye conditions. Four minor diseases with smaller representation (retinal detachment, hemorrhage, nevus and hypertensive retinopathy) are incorporated into the “Other” class.

| Condition | Accuracy | Sensitivity | Specificity | PPV | NPV | AUC | F1 |
| --- | --- | --- | --- | --- | --- | --- | --- |
| diabetic retinopathy | 0.974 | 0.746 | 0.99 | 0.846 | 0.982 | 0.981 | **0.793** |
| macular edema | 0.986 | 0.648 | 0.995 | 0.792 | 0.99 | 0.981 | 0.712 |
| scar | 0.99 | 0.786 | 0.994 | 0.688 | 0.996 | 0.969 | 0.733 |
| amd | 0.974 | 0.671 | 0.981 | 0.482 | 0.991 | 0.96 | 0.561 |
| vascular occlusion | 0.989 | 0.636 | 0.992 | 0.341 | 0.998 | 0.906 | 0.444 |
| drusens | 0.895 | 0.748 | 0.926 | 0.678 | 0.946 | 0.929 | 0.711 |
| myopic fundus | **0.993** | 0.63 | 0.998 | 0.829 | 0.995 | **0.993** | 0.716 |
| increased cup disc | 0.891 | 0.734 | 0.929 | 0.713 | 0.936 | 0.936 | 0.724 |
| other | 0.931 | 0.358 | 0.956 | 0.263 | 0.971 | 0.78 | 0.303 |

Note that at the level of these scores, the effect of this manipulation is negligible. We experimented with a *weighted* loss function where the penalty assigned to a class is inversely proportional its frequency in the training set. We also ran this same experiment on the reduced number of classes. These additions did not improve the performance of the model.

Another common approach to dealing with class imbalance is to use *ensemble methods*, whereby the outputs of various trained models are combined. The downside to such approaches is the additional computational cost (in both training and clinical application in healthcare institutions). We train three models, one with over-sampling in the “Other” class, and another with over-sampling in three classes (nevus, hypertensive retinopathy and hemorrhage), and one without any sampling effects. To explore the benefits of an ensemble approach, we search a 3-dimensional parameter space of weights with which we combine the outputs of these three models. In Table 7 we present the results of the best-performing ensemble model, whose weights are

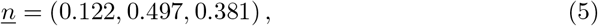

respectively.

**Table 7.** Performance metrics for various eye conditions for an ensemble method combining three separate models.

| Condition | Accuracy | Sensitivity | Specificity | PPV | NPV | AUC | F1 |
| --- | --- | --- | --- | --- | --- | --- | --- |
| diabetic_retinopathy | 0.975 | 0.77 | 0.989 | 0.837 | 0.984 | 0.985 | **0.802** |
| macular_edema | 0.986 | 0.67 | 0.995 | 0.787 | 0.991 | 0.99 | 0.724 |
| scar | 0.99 | 0.554 | 0.997 | 0.795 | 0.992 | 0.956 | 0.653 |
| nevus | 0.876 | 0.207 | 0.882 | 0.016 | 0.992 | 0.662 | 0.029 |
| amd | 0.97 | 0.707 | 0.976 | 0.436 | 0.992 | 0.956 | 0.54 |
| vascular_occlusion | 0.993 | 0.727 | 0.994 | 0.471 | 0.998 | 0.96 | 0.571 |
| hypertensive_retinopathy | 0.978 | 0.186 | 0.993 | 0.324 | 0.985 | 0.821 | 0.237 |
| drusens | 0.904 | 0.734 | 0.939 | 0.715 | 0.944 | 0.93 | 0.724 |
| hemorrhage | 0.99 | 0.647 | 0.991 | 0.282 | 0.998 | 0.912 | 0.393 |
| retinal_detachment | 0.0 | 1.0 | 0.0 | 0.0 | 0.0 | 0.969 | 0.001 |
| myopic_fundus | **0.994** | 0.674 | 0.998 | 0.838 | 0.995 | **0.994** | 0.747 |
| increased_cup_disc | 0.903 | 0.789 | 0.93 | 0.729 | 0.948 | 0.949 | 0.758 |
| other | 0.936 | 0.547 | 0.953 | 0.338 | 0.98 | 0.875 | 0.418 |

#### 3.2.3 Sensitivity Analysis

We also ran the simple baseline models on two different training sets (one entirely Canon, and the other entirely Nikon). These models were then tested on images solely of the other camera type. These results are collated in Tables 8 and 9 respectively. It is clear that the model trained on Canon images performs substantially better than its Nikon counterpart.

**Table 8.** Performance metrics for various eye conditions. Trained on Canon images, tested on Nikon images.

| Condition | Accuracy | Sensitivity | Specificity | PPV | NPV | F1 |
| --- | --- | --- | --- | --- | --- | --- |
| diabetic retinopathy | 0.977 | 0.779 | 0.991 | 0.858 | 0.984 | **0.817** |
| macular edema | 0.989 | 0.843 | 0.993 | 0.713 | 0.997 | 0.773 |
| scar | 0.986 | 0.766 | 0.989 | 0.545 | 0.996 | 0.637 |
| nevus | 0.007 | 1.0 | 0.0 | 0.007 |  | 0.013 |
| amd | 0.968 | 0.656 | 0.973 | 0.285 | 0.994 | 0.397 |
| vascular occlusion | **0.997** | 0.423 | 0.999 | 0.786 | 0.997 | 0.55 |
| hypertensive retinopathy | 0.979 | 0.093 | 0.994 | 0.22 | 0.984 | 0.13 |
| drusens | 0.901 | 0.611 | 0.956 | 0.723 | 0.929 | 0.662 |
| hemorrhage | 0.995 | 0.25 | 0.999 | 0.429 | 0.997 | 0.316 |
| retinal detachment | 0.0 | 1.0 | 0.0 | 0.0 |  | 0.0 |
| myopic fundus | 0.992 | 0.617 | 0.996 | 0.627 | 0.996 | 0.622 |
| increased cup disc | 0.906 | 0.695 | 0.938 | 0.628 | 0.953 | 0.66 |
| other | 0.92 | 0.375 | 0.945 | 0.242 | 0.97 | 0.295 |

**Table 9.** Performance metrics for various eye conditions. Model trained on Nikon images, tested on Canon images.

| Condition | Accuracy | Sensitivity | Specificity | PPV | NPV | F1 |
| --- | --- | --- | --- | --- | --- | --- |
| diabetic retinopathy | 0.963 | 0.682 | 0.981 | 0.71 | 0.979 | 0.696 |
| macular edema | 0.983 | 0.605 | 0.993 | 0.702 | 0.989 | 0.65 |
| scar | 0.984 | 0.495 | 0.993 | 0.577 | 0.991 | 0.533 |
| nevus | 0.849 | 0.229 | 0.855 | 0.014 | 0.992 | 0.027 |
| amd | 0.96 | 0.351 | 0.976 | 0.283 | 0.983 | 0.313 |
| vascular occlusion | **0.992** | 0.338 | 0.996 | 0.4 | 0.995 | 0.366 |
| hypertensive retinopathy | 0.972 | 0.161 | 0.986 | 0.175 | 0.985 | 0.168 |
| drusens | 0.882 | 0.63 | 0.937 | 0.686 | 0.92 | 0.657 |
| hemorrhage | 0.989 | 0.097 | 0.995 | 0.127 | 0.994 | 0.11 |
| retinal detachment | 0.001 | 1.0 | 0.0 | 0.001 | 0.001 | 0.001 |
| myopic fundus | 0.976 | 0.678 | 0.982 | 0.423 | 0.993 | 0.521 |
| increased cup disc | 0.867 | 0.722 | 0.912 | 0.712 | 0.915 | **0.717** |
| other | 0.838 | 0.446 | 0.858 | 0.135 | 0.969 | 0.208 |

#### 3.2.4 Image Quality Analysis

To study the effects of image quality, we ran an experiment using on images classified as “Adequate” based on the metrics described in Section 2.1. The results of this baseline are in Table 10.

**Table 10.** Performance metrics for various eye conditions, trained and tested on “Adequate” quality images only.

| Condition | Accuracy | Sensitivity | Specificity | PPV | NPV | AUC | F1 |
| --- | --- | --- | --- | --- | --- | --- | --- |
| diabetic retinopathy | 0.977 | 0.754 | 0.992 | 0.862 | 0.983 | 0.98 | **0.805** |
| macular edema | 0.989 | 0.719 | 0.995 | 0.767 | 0.994 | 0.994 | 0.742 |
| scar | 0.987 | 0.772 | 0.992 | 0.657 | 0.995 | 0.965 | 0.71 |
| nevus | 0.778 | 0.381 | 0.781 | 0.013 | 0.994 | 0.623 | 0.025 |
| amd | 0.971 | 0.657 | 0.978 | 0.419 | 0.992 | 0.965 | 0.512 |
| vascular occlusion | **0.993** | 0.579 | 0.996 | 0.478 | 0.997 | 0.91 | 0.524 |
| hypertensive retinopathy | 0.975 | 0.218 | 0.99 | 0.3 | 0.985 | 0.788 | 0.253 |
| drusens | 0.889 | 0.748 | 0.919 | 0.666 | 0.944 | 0.91 | 0.704 |
| hemorrhage | 0.984 | 0.333 | 0.987 | 0.098 | 0.997 | 0.724 | 0.151 |
| retinal detachment | 0.0 | 1.0 | 0.0 | 0.0 | 0.0 | 0.66 | 0.001 |
| myopic fundus | **0.993** | 0.744 | 0.997 | 0.78 | 0.996 | **0.995** | 0.762 |
| increased cup disc | 0.897 | 0.825 | 0.915 | 0.702 | 0.955 | 0.945 | 0.759 |
| other | 0.939 | 0.342 | 0.965 | 0.301 | 0.971 | 0.819 | 0.32 |

## 4 Discussion

The goal of this work was to provide a proof-of-principle model on a new dataset of retinal fundus images. We demonstrate that a ResNet-50 model, with appropriate augmentation and class imbalance strategies, offers strong classification performance across a large number of diseases. We propose that this model could be used as a first-order diagnosing tool, in tandem with clinical specialists. This is of particular importance in LMICs where the ability to reduce the work-load of a limited number of ophthalmologists is essential.

In this work we have performed a considerable number of experiments to determine the best performing model. These included experimenting with number of epochs, batch size, regularization methods, class imbalance techniques, image quality, and data augmentation.

We experimented with batch sizes 2, 4, 8, 16, 32 and found that a batch size of 8 consistently out-performed the others. With our choice of model and relatively small dataset size, after trying various epochs we observed that 6 epochs was optimal. We attempted various regularization schemes, including dropout and L2-regularization to probe a higher number epochs but did not observe meaningful improvement. The outcome of this experimentation is the model whose performance is detailed in Table 4.

Notice that there is considerable variation in the model’s performance with respect to each individual disease. Broadly speaking, those classes with larger representation in the training set are more easily identified by our model. In particular, the baseline model performs well on diabetic retinopathy, macular edema, scar, drusens, myopic fundus and increased cup disc ratio in particular.

Expectedly, diseases such as nondiabetic retinal hemorrhage, retinal detachment and nevus have poor performance: each of these represent less than 1% of the dataset. In particular, there are only 7 instances of retinal detachment in the dataset. Interestingly, the model also struggles to identify hypertensive retinopathy despite the fact that it is represented in the dataset more frequently than some better performing classes.

The class “Other” is also not easily identified by our baseline model. We speculate that this is because this class contains a variety of different diseases which may present in highly unique ways. This diversity of features within one class could present difficulties for a model to generalize. In future work it would be interesting to obtain finer-grained clinical labelling of these images in order to train a specialized model on these smaller classes.

### Class Imbalance

To try to address the class imbalance, we performed various experiments. In Table 5 we present the results for an experiment where 5000 “Normal” labelled images were removed - thus addressing the imbalance between images with disease and without. This model presents a marginal improvement on our original baseline in 4, with 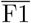 improving from 0.68 to 0.70.

In Table 6 we reduce the size of the search space by consolidating four of the under-represented diseases within the “Other” class. As above, this only generates a minor improvement upon the baseline.

In Table 7 we experimented with an ensemble approach. We observe small improvement in performance in some poorer-performing classes (for example, there is small improvement in the “Other” F1-score). However, we conclude that these small gains do not justify the increased computational cost and complexity of this model.

### Image Quality

We chose to include images of all quality in the training phase to make our model more robust. However, in Table 10 we demonstrate that restricting training/testing to “Adequate” images clearly leads to improved performance. This of-course comes with a reduction in generalizability to real-world, potentially lower quality, images. We choose to prioritise robustness to a variety of image qualities in real-world applications, rather than improved performance on a subset of images. With this in mind, we also chose not to do any further image pre-processing with regards to blurriness, blemishes, exposure etc. In effect, we kept the natural noise of the training set to improve the model’s ability to be useful in real-world scenarios.

### Comparison with Literature

There are several works in the literature on retinal disease classification using computer vision techniques. To our knowledge, our work is the first model trained on the BREST dataset. Our work provides an example of using these techniques on images obtained from patients from a part of the world which is under-represented in the literature. It is our contention that this work could be used in tandem with others to provide a more robust and generally applicable diagnosis tool. Our work is a model trained on a unique set of labels: existing models in the literature are trained to detect different sets of retinal diseases.

Here we collect a small number of works in this area to demonstrate where our work sits in the body of literature. In [14] the authors classify five increasing gradings of diabetic retinopathy by training a model based on a highly imbalanced dataset. In [16] the authors classified 39 retinal fundus diseases by training on a dataset with over 250,000 labels collected from across the United States, India and China. In [17] the authors detect 8 different types of fundus legions using a Chinese dataset.

The power of our work is two-fold. Firstly, we train our model on an entirely new dataset, widening the access of this type of work to a new demographic. Secondly, our model is trained to identify a unique set of retinal diseases. We propose that in future work an ensemble approach involving various multi-label classifiers could provide a powerful general diagnosis tool.

### Comments

While we have demonstrated the efficacy of our model in managing multilabel classification, it remains essential to contemplate whether a disease-specific model is more fitting for certain diseases or each disease individually. For instance, if the model’s objective is to provide support to patients with a particular disease or a combination of diseases, predict the risk of a specific disease, or recommend a particular treatment pathway, then employing personalized models trained separately for each class might be the optimal choice. Nevertheless, employing multiple models can be computationally intensive, presenting challenges for healthcare institutions. In such scenarios, opting for a more generalized model, such as the multilabel framework presented here, would be advantageous. This approach provides an efficient initial classification, which could facilitate rapid triaging and screening.

Multi-label classification involves predicting multiple labels for each instance, making the task inherently more complex than binary or multi-class classification. As the number of labels increases, the feature space grows exponentially, leading to the “curse of dimensionality”. This can affect model training efficiency and performance. Thus, models trained on one multi-label dataset may not generalize well to other datasets due to differences in label semantics, distributions, or dependencies. As we discussed above, some labels may have significantly fewer instances than others, leading to imbalances that can affect model training and performance.

## 5 Conclusions

In this work we have demonstrated the applicability of computer vision techniques to the problem of retinal disease diagnosis. To the best of our knowledge, this work is the first baseline model trained on the BREST dataset [18].

In our work we have proven the efficacy of the ResNet-50 model in a multi-label retinal disease classification. We experimented with hyperparameters, augmentation and class imbalance methods to arrive at the best performing model.

In future works it would be interesting to build upon this baseline with more complicated architectures. It would also be instructive to combine this model with other retinal fundus multi-label models (trained on a variety of datasets to identify *different* sets of diseases) to construct a model with even greater applicability.

## Data Availability

The dataset is available on PhysioNet at https://physionet.org/content/brazilian-ophthalmological/1.0.0/. In order to access the data, users are required to be a credentialed user and complete various training modules on the subject of data ethics, fair-use and safety.

## Declarations

### Ethics Approval

This project was approved by the São Paulo Federal University - UNIFESP institutional review board (CAAE 33842220.7.0000.5505). The requirement for individual consent was waived. In this dataset, all images were anonymized with all identifiable patient information removed.

## Acknowledgements

The creation of this dataset project was funded by Instituto da Visão - IPEPO and Lemann Foundation. The authors would like to acknowledge the use of the University of Oxford Advanced Research Computing (ARC) facility in carrying out this work. http://dx.doi.org/10.5281/zenodo.22558

## Author Contributions

DG and JY wrote the initial draft of the manuscript. DG and JY set up the code. JY conceived the study. DG ran the experiments. All authors revised the manuscript.

## Data Availability

The dataset is available on PhysioNet [19] at https://physionet.org/content/brazilian-ophthalmological/1.0.0/. In order to access the data, users are required to be a credentialed user and complete various training modules on the subject of data ethics, fair-use and safety.

## Code Availability

Code and supplementary information for this paper are available alongside publication, and can be found at https://github.com/dewigould/BRESTMultiLabelOpthalmology.

### Appendix A Supplementary Results

In Table A1 we collect the results for a baseline test with batch size 8, where the thresholds are *not* optimized:

**Supplementary Table A1.** Performance metrics for various eye conditions. 6 epochs, batch size 8. This model has 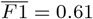 and 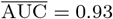.

| Condition | Accuracy | Sensitivity | Specificity | PPV | NPV | AUC | F1 |
| --- | --- | --- | --- | --- | --- | --- | --- |
| diabetic retinopathy | 0.976 | 0.784 | 0.989 | 0.839 | 0.985 | 0.979 | **0.811** |
| macular edema | 0.983 | 0.705 | 0.991 | 0.689 | 0.992 | 0.985 | 0.697 |
| scar | 0.984 | 0.732 | 0.988 | 0.526 | 0.995 | 0.976 | 0.612 |
| nevus | 0.991 | 0.0 | 1.0 |  | 0.991 | 0.711 | 0.0 |
| amd | 0.977 | 0.512 | 0.989 | 0.538 | 0.987 | 0.962 | 0.525 |
| vascular occlusion | 0.991 | 0.455 | 0.994 | 0.357 | 0.996 | 0.907 | 0.4 |
| hypertensive retinopathy | 0.978 | 0.186 | 0.993 | 0.324 | 0.985 | 0.827 | 0.237 |
| drusens | 0.895 | 0.462 | 0.986 | 0.869 | 0.898 | 0.919 | 0.604 |
| hemorrhage | **0.993** | 0.118 | 0.998 | 0.222 | 0.995 | 0.78 | 0.154 |
| retinal detachment | 1.0 | 0.0 | 1.0 |  | 1.0 | 0.112 | 0.0 |
| myopic fundus | 0.992 | 0.609 | 0.997 | 0.757 | 0.994 | **0.992** | 0.675 |
| increased cup disc | 0.895 | 0.757 | 0.928 | 0.715 | 0.941 | 0.945 | 0.735 |
| other | 0.947 | 0.197 | 0.979 | 0.297 | 0.965 | 0.832 | 0.237 |

In Table A2 we collect the results for a baseline test with batch size 8, with threshold values chosen to maximise individual F1 scores.

**Supplementary Table A2.**
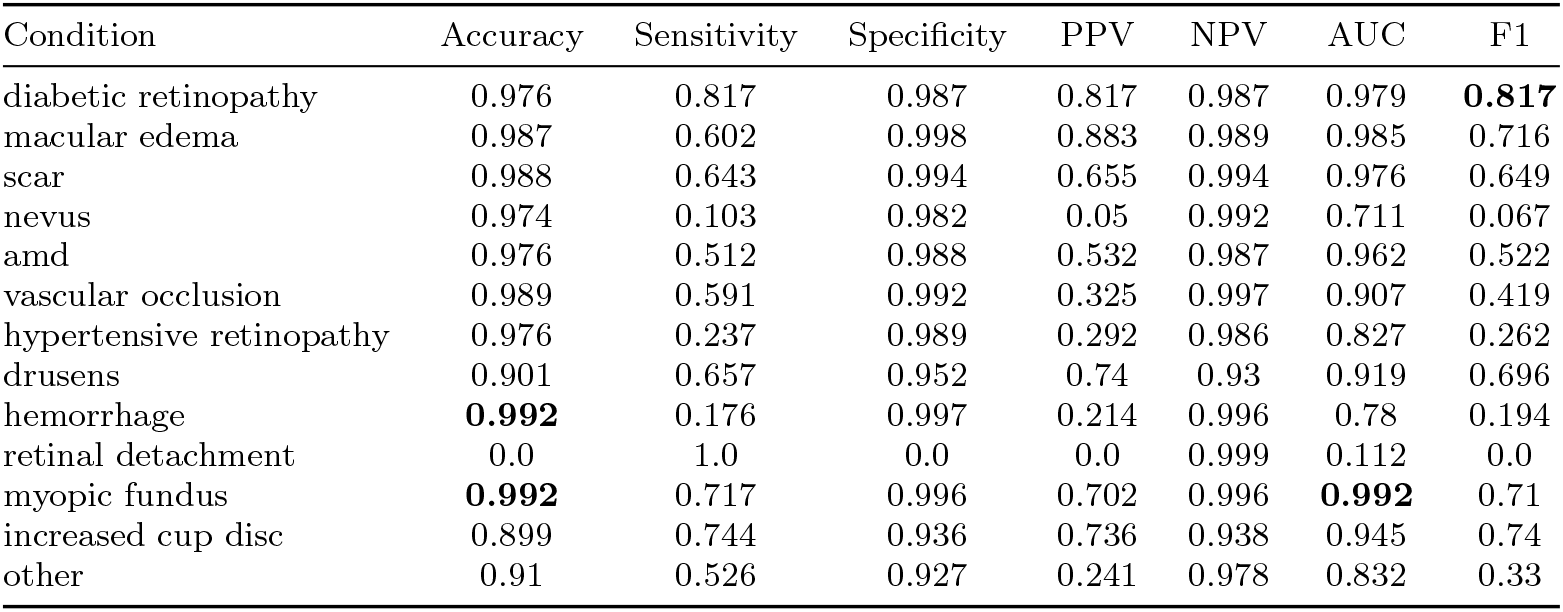
Performance metrics for various eye conditions. 6 epochs, batch size 8. This model has 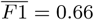 and 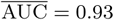.

| Condition | Accuracy | Sensitivity | Specificity | PPV | NPV | AUC | F1 |
| --- | --- | --- | --- | --- | --- | --- | --- |
| diabetic retinopathy | 0.976 | 0.817 | 0.987 | 0.817 | 0.987 | 0.979 | **0.817** |
| macular edema | 0.987 | 0.602 | 0.998 | 0.883 | 0.989 | 0.985 | 0.716 |
| scar | 0.988 | 0.643 | 0.994 | 0.655 | 0.994 | 0.976 | 0.649 |
| nevus | 0.974 | 0.103 | 0.982 | 0.05 | 0.992 | 0.711 | 0.067 |
| amd | 0.976 | 0.512 | 0.988 | 0.532 | 0.987 | 0.962 | 0.522 |
| vascular occlusion | 0.989 | 0.591 | 0.992 | 0.325 | 0.997 | 0.907 | 0.419 |
| hypertensive retinopathy | 0.976 | 0.237 | 0.989 | 0.292 | 0.986 | 0.827 | 0.262 |
| drusens | 0.901 | 0.657 | 0.952 | 0.74 | 0.93 | 0.919 | 0.696 |
| hemorrhage | **0.992** | 0.176 | 0.997 | 0.214 | 0.996 | 0.78 | 0.194 |
| retinal detachment | 0.0 | 1.0 | 0.0 | 0.0 | 0.999 | 0.112 | 0.0 |
| myopic fundus | **0.992** | 0.717 | 0.996 | 0.702 | 0.996 | **0.992** | 0.71 |
| increased cup disc | 0.899 | 0.744 | 0.936 | 0.736 | 0.938 | 0.945 | 0.74 |
| other | 0.91 | 0.526 | 0.927 | 0.241 | 0.978 | 0.832 | 0.33 |

## Footnotes

1 This number is most often 13, the number of disease classes in Table 1. However, occasionally this will be less than 13 when we experiment with methods to account for class imbalance.

